# Protein aggregation of DISC1, as assayed by insolubility, varies across the brain of an individual with schizophrenia and Alzheimer’s disease

**DOI:** 10.1101/2023.08.01.23293413

**Authors:** Bobana Samardžija, Éva Renner, Miklós Palkovits, Nicholas J. Bradshaw

## Abstract

**Objective:** Subgroups of mental illness patients have been seen to display disturbed proteostasis, with specific proteins aggregating in their brain, which is generally determined by assaying protein insolubility in the *post mortem* samples. Such studies typically only look at one region of the brain, and therefore we aimed to determine the distribution of protein across a single brain, using this insolubility-based approach.

**Methods:** We looked at 20 post *mortem* tissue samples from across the brain of a single patient, with schizophrenia and Alzheimer’s disease, determined which protein(s) aggregated in his brain relative to controls, based on purification of insoluble protein fractions. The individual samples were then similarly analysed.

**Results:** Disrupted in Schizophrenia 1 (DISC1) protein was seen to be insoluble in the patient’s brain, however in a very heterogenous picture, with differences in insoluble DISC1 even between samples of the same region, but opposite hemispheres.

**Conclusions:** While caution must be taken in extrapolating from a single individual, this raises the possibility that aggregates of DISC1 may spread throughout the brain, as is the case for proteins in neurodegenerative disorders, and suggests that current studies looking at single brain regions may be underestimating the prevalence of protein aggregates in schizophrenia.

## Introduction

An emerging field of schizophrenia research, is to consider it as a disease partially characterised by disrupted proteostasis, and in particular investigating whether specific protein may form aggregates in the brains of patients, in partial analogy to similar phenomena in neurodegenerative diseases (Bradshaw & Korth 2019). To date, a number of different proteins have been implicated as aggregating in schizophrenia, including DISC1 (Disrupted in Schizophrenia 1), CRMP1 (Collapsin Response Mediator Protein 1), TRIOBP (TRIO & F-actin Binding Protein) and NPAS3 (Neuronal PAS domain containing protein 3) (Leliveld *et al*. 2008; Bader *et al*. 2012; Bradshaw *et al*. 2014; Nucifora *et al*. 2016).

The main method used to identify or confirm aggregation of these proteins is through the acquisition of *post mortem* tissue samples from the brains of patients and controls, homogenising them, and then purifying the insoluble protein fraction from them. While not all proteins that are insoluble are aggregates, if a normally soluble protein appears in the insoluble protein fraction of an individual, this is likely because that protein is misfolding or aggregating in this specific individual (Bradshaw & Korth 2019). This approach has been used on a number of different schizophrenia cohorts to identify both specific and general protein aggregation (table 1). Notably, however, in these studies normally only one brain region has been looked at in each individual, due to the scarcity of well-preserved brain tissue from schizophrenia patients, and the region of the brain used varies between studies (table 1).

**Table 1:** Regions of the brain used in previous studies of the insoluble protein in major mental illness, along with details of the diagnoses investigated (not including control individuals) in each, and the proteins investigated. Key to diagnoses: SCZ: schizophrenia, BPD: bipolar disorder, MDD: major depressive disorder. “Total protein” indicates the use of proteomics-based approaches for protein identification and/or looking at total insoluble protein levels, while listing of specific protein names indicates Western blotting for hypothesis-based tests.

| Brain region | Diagnoses | Main protein studied | Publication |
| --- | --- | --- | --- |
| Anterior cingulate cortex (BA23) | SCZ, BPD, MDD | DISC1 | (Leliveld *et al.* 2008) |
| Anterior cingulate cortex (BA23) | SCZ, BPD, MDD | Dysbindin-1 | (Ottis *et al.* 2011) |
| Anterior cingulate cortex (BA23) | SCZ, BPD, MDD | CRMP1 | (Bader *et al.* 2012) |
| Anterior cingulate cortex (BA23) | SCZ, MDD | TRIOBP | (Zaharija *et al.* 2022) |
| Insular cortex | Suicide, MDD | NPAS3 | (Samardžija *et al.* 2021) |
| Insular cortex | Suicide, MDD | DISC1, CRMP1, TRIOBP | (Samardžija *et al.* 2023) |
| Prefrontal cortex (BA10) | SCZ | Total protein | (Nucifora *et al.* 2019) |
| Prefrontal cortex (BA46) | SCZ | Total protein | (Nucifora *et al.* 2019) |
| Superior temporal gyrus | SCZ | Total protein | (Nucifora *et al.* 2019) |

It is thus difficult to directly compare results of these studies, as it is unclear whether aggregation of these proteins is consistent across the brain, or shows regional variations. Here we therefore examine protein aggregation across 20 brain regions of a single individual, 10 per hemisphere.

## Materials & Methods

### Brain samples

Samples of human brain tissue were collected as part of the Lenhossék Program, a country-wide initiative in Hungary, after obtaining familial consent or legal permission. Within six hours of death, brain samples were extracted using a micropunch technique (Palkovits 1973; Palkovits 1985), snap frozen in liquid nitrogen, and stored at -80 °C until required.

Samples used in this study came from an individual, patient R (arbitrary designation given without knowledge of the patient’s identity), diagnosed with schizophrenia and Alzheimer’s disease. Samples were also collected from three individuals with Alzheimer’s disease and three control individuals, from at least 4 of the following 5 regions: the frontal cortex, lateral orbitofrontal gyrus, the occipital cortex (area striata), somatomotor cortex (precentral gyrus) and the somatosensory cortex (postcentral gyrus). Samples from patient R were from the same regions and others (table 2), with two samples per region: one from each hemisphere of the brain.

**Table 2:** Brain regions investigated from patient R in this study. “Labels” are used in figure 2. Unless samples from the left hemisphere directly correspond to samples in the right hemisphere

| Label | Region name | Brodmann area |
| --- | --- | --- |
| 1 | Dorsal premotor cortex | BA6a |
| 2 | Frontal cortex |  |
| 3 | Fusiform gyrus | BA36, BA37 |
| 4 | Lateral orbitofrontal gyrus |  |
| 5 | Middle frontal gyrus | BA9 |
| 6 | Occipital cortex, area striata | BA17 |
| 7 | Parietal cortex |  |
| 8 | Somatomotor cortex (precentral gyrus) | BA4 (left), BA3 (right) |
| 9 | Superior frontal gyrus |  |
| 10 | Temporal cortex |  |

### Insoluble protein fraction purification

All laboratory work on the samples was performed by a researcher who was blinded as to the nature of each sample, which were decoded only after completion of Western blotting. Brain samples were homogenised to 10% w/v in VRL buffer (50 mM HEPES / 250 mM sucrose / 5 mM magnesium chloride / 100 mM potassium acetate) containing 2 mM PMSF plus protease inhibitor cocktail. 400 μl of each sample then had its insoluble protein fraction purified using an established protocol (original protocol: Leliveld *et al*. 2008; most recent iteraction: Zaharija *et al*. 2022) in which insoluble proteins are repeatedly suspended in solubilisation buffers (variously containing high salt, high sucrose and detergents), and then have their insoluble pellet isolated by ultracentrifugation (Sorvall MTX150 with S140-AT rotor, Thermo Fisher Scientific). Both samples of the original non-fractionated homogenate, and the purified final insoluble pellet were then analysed by blotting.

### Western blotting

Samples were denatured in Laemmeli sample buffer containing DTT and then run on SDS-PAGE gels. Gels were transferred to PVDF membranes (Macherey-Nagel) using a Transblot Turbo system (Bio-Rad). Membranes were blocked in PBS-T (PBS / 0.05% Tween-20) mixed with 5% milk powder, and then stained with primary and secondary antibodies in PBS-T, with washes in between. Signal was visualised using ECL (Thermo Fisher Scientific) on ChemiDoc MP Imaging System and associated software (Bio-Rad). Antibodies were purchased from Atlas Antibodies (anti-TRIOBP, HPA019769), Origene (anti-β-actin, OG-TA811000), PromoCell (anti-NPAS3 PK-AB718-4107), ProSci (anti-CRMP1 3625) and Thermo Fisher Scientific (anti-DISC1 40-6800, secondary antibodies 31430 & 656120).

## Results

Patient R was a male diagnosed with both schizophrenia and Alzheimer’s disease, who died in his late 70s. His brain was generously donated to the Lenhossék Program, and 20 tissue samples were available for this study, representing 10 regions from each hemisphere. Each of these samples had their insoluble protein fraction purified, according to previously described and validated protocols (Zaharija *et al*. 2022), as this fraction would be expected to contain any aggregating proteins.

Insoluble material from 5 of the brain regions was pooled, and combined to equivalent insoluble protein fractions from three control individuals and three patients with Alzheimer’s disease. Patient R showed clear indication of insoluble DISC1, as did one other Alzheimer’s disease patient, compared to the other samples (figure 1). Specificity of the DISC1 antibody was previously validated by knockdown (Marley & von Zastrow 2010). For the other proteins examined, CRMP1, NPAS3 or TRIOBP, there was no clear enrichment of insoluble protein in patient R compared to the other samples examined (figure 1A). It therefore appears likely that patient R had DISC1 aggregation in his brain.

**Figure 1:**
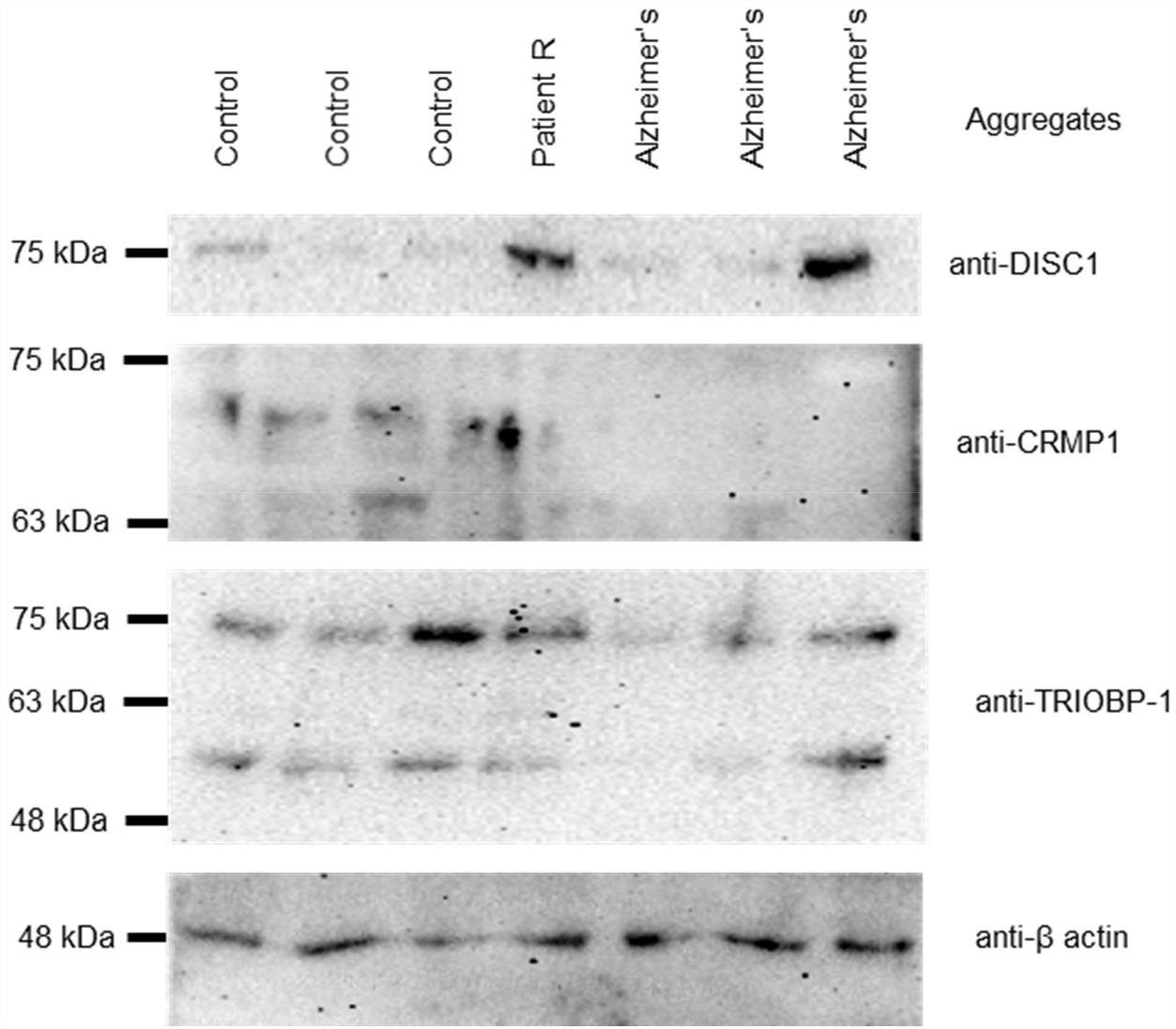
(A) Presence of proteins in the purified insoluble fractions of mixed brain homogenates from 7 individuals: 3 control individuals, patient R, who had schizophrenia and Alzheimer’s disease, and 3 individuals with Alzheimer’s disease. Of these, insoluble DISC1 showed the highest prominence in patient R, compared to the control individuals.

Next, all 20 samples from patient R were investigated by Western blotting individually, to determine whether this insoluble DISC1 was evenly distributed throughout the brain. Strikingly, while some immunoreactivity was seen in purified insoluble pellets in most brain regions (with the exception of the occipital, somatomotor and somatosensory cortices), the strength of the signal varied between regions (figure 2A). This variation in insoluble protein did not correlate with variation in total levels of DISC1 in the original brain region homogenate (figure 2B). For most of the brain regions covered, brain tissue was used from both hemispheres, revealing notable differences between them: the left hemisphere showed most insoluble DISC1 in the dorsal premotor cortex and temporal cortex, while the right showed most in the fusiform gyrus and parietal cortex (figure 2C,D). While this only represents the situation in a single patient, it suggests that DISC1 aggregation is not uniform across the brains of patients in which it manifests.

**Figure 2:**
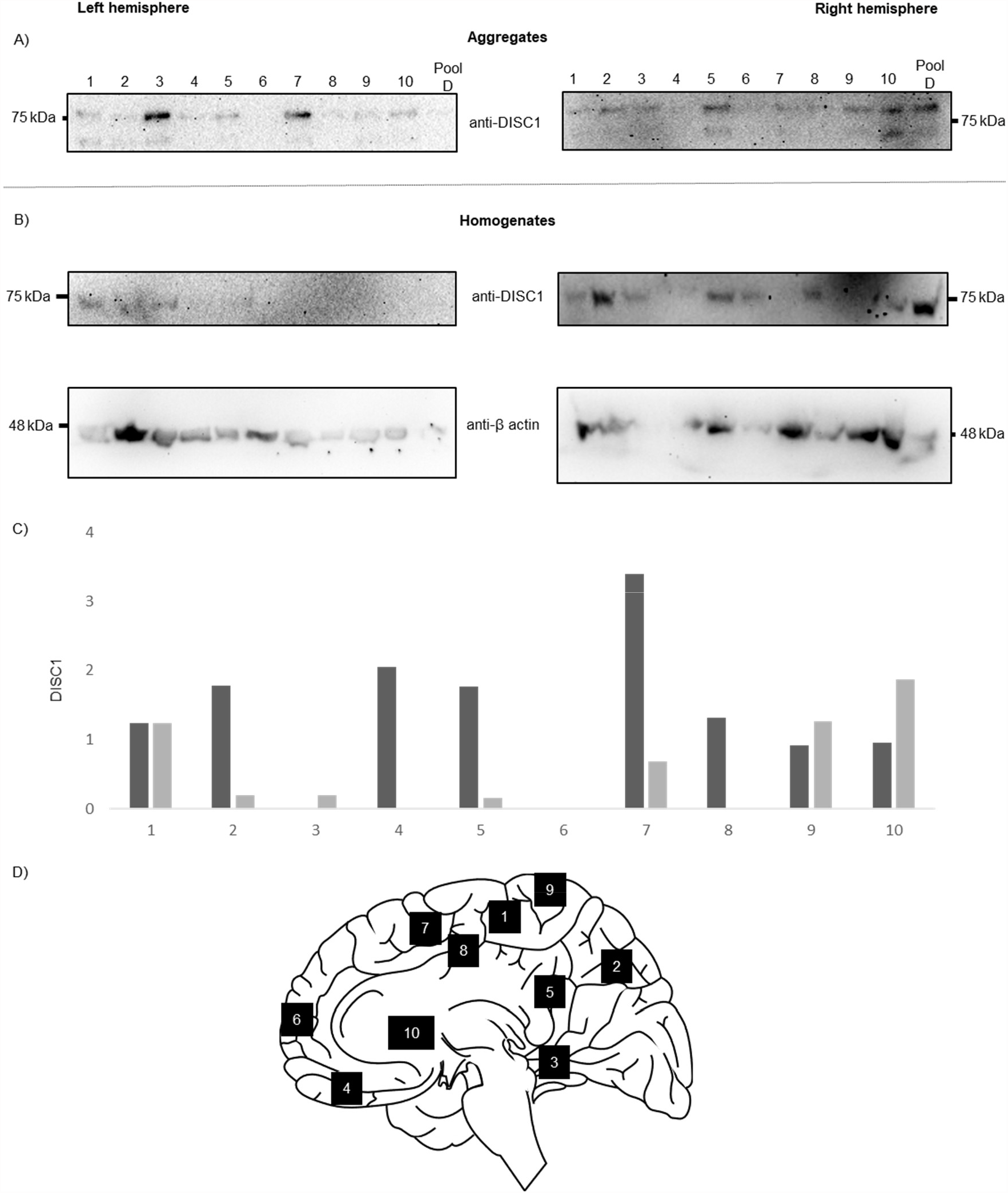
(A) Presence of DISC1 in the purified insoluble fraction of 20 brain samples: all taken from patient R, and representing left and right hemisphere samples of different brain regions. Key to brain region numbers is given in table 2. (B) DISC1 in the original, non-purified, brain homogenates. (C) Quantified levels of DISC1 in the insoluble brain fractions, corrected for the level of actin in the original homogenate. Darker bars indicate signal in the left hemisphere, lighter bars indicate signal in the right hemisphere. (D) Approximate locations that the samples were taken from. Sagittal brain image by Stoyo Karamihalev, scidraw.io/drawing/395, doi:10.5281/zenodo.4312476.

## Discussion

For this study, aggregation of DISC1 and other proteins was defined based on their insolubility, on the premise that if a normally soluble protein is insoluble in brain tissue of specific patients, then this is indicative of the protein aggregating (Bradshaw & Korth 2019). This technique has been used for over 15 years (Leliveld *et al*. 2008), and has been refined and optimised over time to ensure consistency of findings when the assay is performed by the same individual (Zaharija *et al*. 2022), while in cell culture models of DISC1 and other schizophrenia-related proteins correlates well with the formation of visible protein aggregates, as detected using immunofluorescent microscopy (Ottis *et al*. 2011; Bader *et al*. 2012; Bradshaw *et al*. 2014).

This approach requires high-quality *post mortem* brain tissue, which has been flash frozen after a relatively short *post mortem* interval, and stored without any fixatives or preservatives. Such material is relatively hard to come by from schizophrenia patients, meaning that studies are often limited in sample numbers and confined to only a single region of the brain. It is therefore imperative to know whether aggregation of DISC1, and other schizophrenia-specific proteins, is homogenous or heterogenous across the brains of patients. Therefore, in this study, we performed insolubility-based assays on multiple brain regions from a single patient. It is established that each of these individual proteins aggregates in only a subset of patients (Leliveld *et al*. 2008; Ottis *et al*. 2011; Bader *et al*. 2012; Zaharija *et al*. 2022; Samardžija *et al*. 2021), therefore a pooled brain sample from this patient was compared with those from control individuals, revealing that he was likely to have aggerated DISC1, but not CRMP1, NPAS3 or TRIOBP-1.

Upon looking at the individual brain regions, it became obvious that there is significant variation in insoluble DISC1 levels, even between the same regions of each hemisphere. This is consistent with the idea that aggregates of DISC1 or other proteins may begin in specific regions of the brain and then spread, as is common in neurodegenerative disorders (Davis *et al*. 2018). Indeed, *in vitro* assays have already shown DISC1 to be cell invasive (Ottis *et al*. 2011; Zhu *et al*. 2017), while exosomes containing dysbindin-1, another protein implicated as aggregating in mental illness (Ottis *et al*. 2011), were cell invasive when injected into a mouse brain (Zhu *et al*. 2015).

Of the brain regions examined, the dorsal premotor cortex and superior frontal gyrus showed fairly consistent levels of insoluble DISC1 in each hemisphere. The frontal cortex, lateral orbifrontal gyrus, middle frontal gyrus, parietal cortex and somatomotor cortex, meanwhile, each showed notably more insoluble DISC1 in the left hemisphere, with only the temporal cortex showing considerably more in the right. Notably, a recent meta-analysis showed many of these regions show grey matter lost in schizophrenia patients compared to controls, some of which are hemisphere specific (Picó-Pérez *et al*. 2022).

Some caution must be taken in generalising these results, given that they are taken from only one individual, who may not be representative of all schizophrenia patients. Similarly, it must also be noted that this individual was diagnosed with Alzheimer’s disease in addition to schizophrenia. While no study has, to the best of our knowledge, directly associated DISC1 aggregation with this condition, it cannot be formally excluded that the pathology of Alzheimer’s disease may affect DISC1 aggregation (or vice versa).

Nevertheless, this emphasises that studies looking at protein aggregation in major mental illness need to consider which locations brain samples are taken from. It also raises the possibility that looking at only one brain region would cause some individuals to be labelled as not having protein aggregation, when in fact it is present elsewhere in the brain, leading to underestimation of the prevalence of protein aggregation. It should therefore be a priority for the field to identify brain regions that are critical for aggregation of DISC1, and other proteins, in schizophrenia.

## Data Availability

All data produced in the present work are contained in the manuscript.

## Acknowledgements

This work was funded by the Alexander von Humboldt Foundation (1142747-HRV-IP), the Croatian Science Foundation (IP-2018-01-9424 and DOK-2020-01-8580), and the Hungarian National Research, Development and Innovation Office (NKFIH 2017-1.2.1-NKP-2017-00002, National Brain Research Program NAP 2.0 and NAP2022-I-4/2022, National Brain Research Program NAP 3.0). We thank Beti Zaharija, Elizabeth Bradshaw and the staff of the Human Brain Tissue Bank for technical assistance.

## Author contributions

All authors contributed to experimental design. BS performed the experiments. ÉR and MP collected and curated the brain samples and associated data. BS and NJB analysed the results. NJB drafted the manuscript, while BS prepared the figures. All authors edited and approved the final version.

## Ethics statement

Ethical approval for collection and distribution of these samples was given by the Committee of Science and Research Ethics of the Ministry of Health of Hungary (No. 6008/8/2002/ETT) and the Semmelweis University Regional Committee of Science and Research Ethics (No. 32/1992/TUKEB). Approval for the use of the samples in these experiments was given by the Ethical Committee of the University of Rijeka, Department of Biotechnology (18.02.2022-Bradshaw).

